# Quantifying new threats to health and biomedical literature integrity from rapidly scaled publications and problematic research

**DOI:** 10.1101/2025.07.07.25331008

**Authors:** Matt Spick, Anthony Onoja, Charlie Harrison, Stefan Stender, Jennifer Byrne, Nophar Geifman

**Affiliations:** School of Health Sciences, Faculty of Health and Medical Sciences, University of Surrey, Guildford, United Kingdom, GU2 7XH; Department of Computer Science, Aberystwyth University, Ceredigion, SY23 3DB, UK; Department of Clinical Biochemistry, Rigshospitalet, Copenhagen University Hospital, Copenhagen, Denmark; School of Medical Sciences, Faculty of Medicine and Health, The University of Sydney, Camperdown, New South Wales, Australia; NSW Health Statewide Biobank, NSW Health Pathology, Camperdown, New South Wales, Australia

## Abstract

**Background and Objectives:** The last three years have seen an explosion in published manuscripts analysing open-access health datasets, in many cases presenting misleading or biologically implausible findings. There is a growing evidence base to suggest that this is due in part to AI-assisted and formulaic workflows, and publishers are responding by discouraging submissions employing open-access health datasets.

**Methods:** Here we employ a scientometric analysis to investigate which datasets have seen publication rates deviate from previous trends, especially where this coincides with changes to author geographical origins and increases in formulaic titles.

**Results:** Across 36 datasets we identify nine showing hallmarks of paper mill exploitation (FAERS, NHANES, UK Biobank, FinnGen, the Global Burden of Disease Study, MIMIC, CHARLS, CDC WONDER, and TriNetX). These nine datasets had, in 2025, a combined publication count of 23,005 indexed in the OpenAlex database. This represents an excess of 11,577 publications above the AutoRegressive Integrated Moving Average (ARIMA) forecast trend, and is a 3.0x fold change on the 7,655 publication count for these nine datasets in 2022. We also identified a notable difference in the fold change for China (4.2x) versus the rest of the world (1.9x) and an increase in formulaic titles.

**Conclusions:** These findings highlight potential risks to research integrity in areas such as public health and drug safety, and especially to the accessibility and interoperability principles central to Open Science and FAIR data practices. We argue that permissive open-access data policies naturally facilitate exploitative workflows, and that these findings add to the case for the safeguarding mechanisms to preserve the goals of Open Science

## Introduction

Generative AI (GenAI) and other automation tools have the potential to transform productivity in biomedical and health research. New technologies can, however, also be exploited in problematic ways, and paper mills (entities which mass-produce manuscripts for purchase) are particularly likely to benefit from such productivity gains as their business model relies on the large-scale authoring of inevitably low quality or completely fabricated manuscripts. [1–3] These issues have been identified previously. Notably, the exploitation of the National Health And Nutrition Examination Survey (NHANES) data resource has been highlighted by targeted analysis of individual manuscripts, [4,5] with problematic research practices including lack of false discovery correction, selective data usage / hypothesising after the results are known (also known as HARKing), and data dredging to maximise manuscript counts irrespective of the plausibility of the findings. [6] Investigating individual publications is time-consuming and can be slow to react to new trends, and so trend-analyses can also be helpful when identifying problematic research practices, especially when resources are scarce. [7] As well as rapid growth in the number of publications citing NHANES, [8] concerning trends have also been identified in works poorly employing two-sample Mendelian randomization applied to openly available GWAS data. [9,10] To our knowledge, however, there has not yet been a systematic attempt to quantify the growth in exploitation across the wider field of open access datasets. This has the potential to be a particular problem for datasets in the field of health and epidemiology, especially those that are FAIR compliant (e.g. with API-supported access), which may be vulnerable to AI-supported and pipeline-driven mass production of papers.

These issues are, of course, not new. Genomics provides a case study of a field that experienced an explosion of results, often lacking in biological plausibility or which could not be reproduced. This was addressed through a number of measures, including much more stringent genome-wide significance thresholds, [11] meta-analyses, [12] pre-registration of study designs and protocols to reduce data dredging, [13,14] and adoption of reporting standards such as STREGA, [15] all of which may be helpful in addressing AI-assisted exploitation of health data sources. Other measures employed in genomics, such as use of large-scale datasets and the promotion of open data, [16] may in contrast not work well in the AI-assisted era, as open-access naturally facilitates the type of data dredging and HARKing that has been seen in the last three years. [17,18]

In this work we attempt to establish the impact of AI-assisted formulaic templates we believe are currently in use, by examining trends in the number of publications published in journals and indexed in OpenAlex. We use these findings to discuss whether existing guidelines from policymakers are futureproofed to deal with these new strategies for the mass production of manuscripts, a crucial issue given the efforts to encourage Open Science and adoption of FAIR Guiding Principles. [19–22] In the worst-case scenario, unchecked exploitation and manipulation of FAIR assets could undermine confidence in publication-based dissemination and lead to researchers turning away from open data altogether.

## Methods

To identify health data sources that might be current targets of exploitation, a search of the OpenAlex database via API using the Entrez module from the Biopython library was conducted (version 1.85) for publications dated between 2014 and 2025, [23] related to a list of 36 health or biomedical databases (Table 1) based on the authors’ collective expertise in the field. Alternative search terms for each dataset were used where appropriate for acronyms, or additional Boolean operators were used where acronyms were associated with other issues (for example the PRIDE database of proteomics mass spectrometry information) and the search strings are detailed in Supplementary Materials, Table S1. An additional query was submitted to obtain combined publication counts for ‘at risk’ datasets, using OR logic across dataset terms with deduplication. As a result, totals of combinations of datasets would be expected to be lower than the arithmetic sum of per-dataset counts. All searches were conducted for Title or Abstract, filtered to show type = article, and were run on 3 January 2026.

**Table 1:**
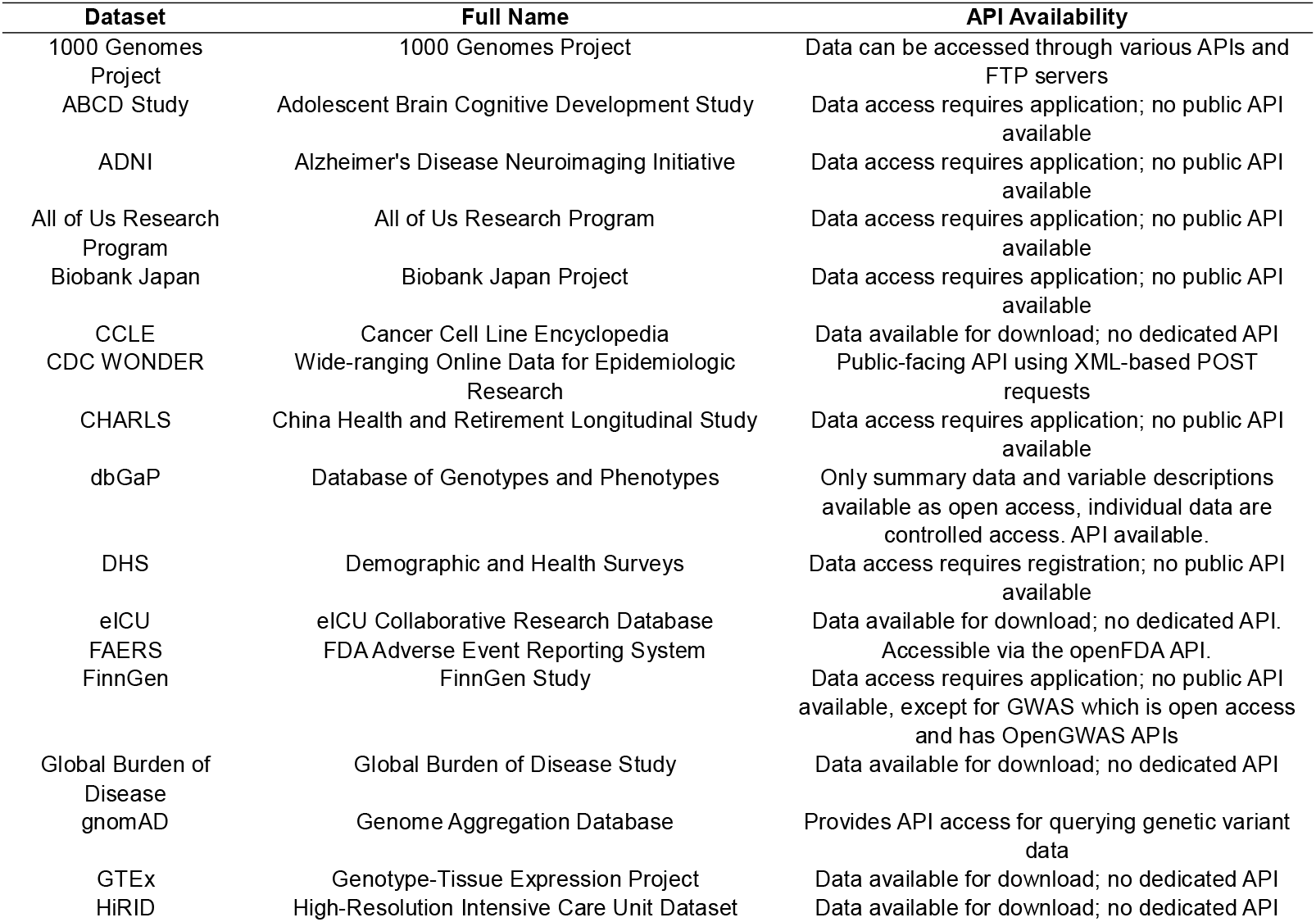
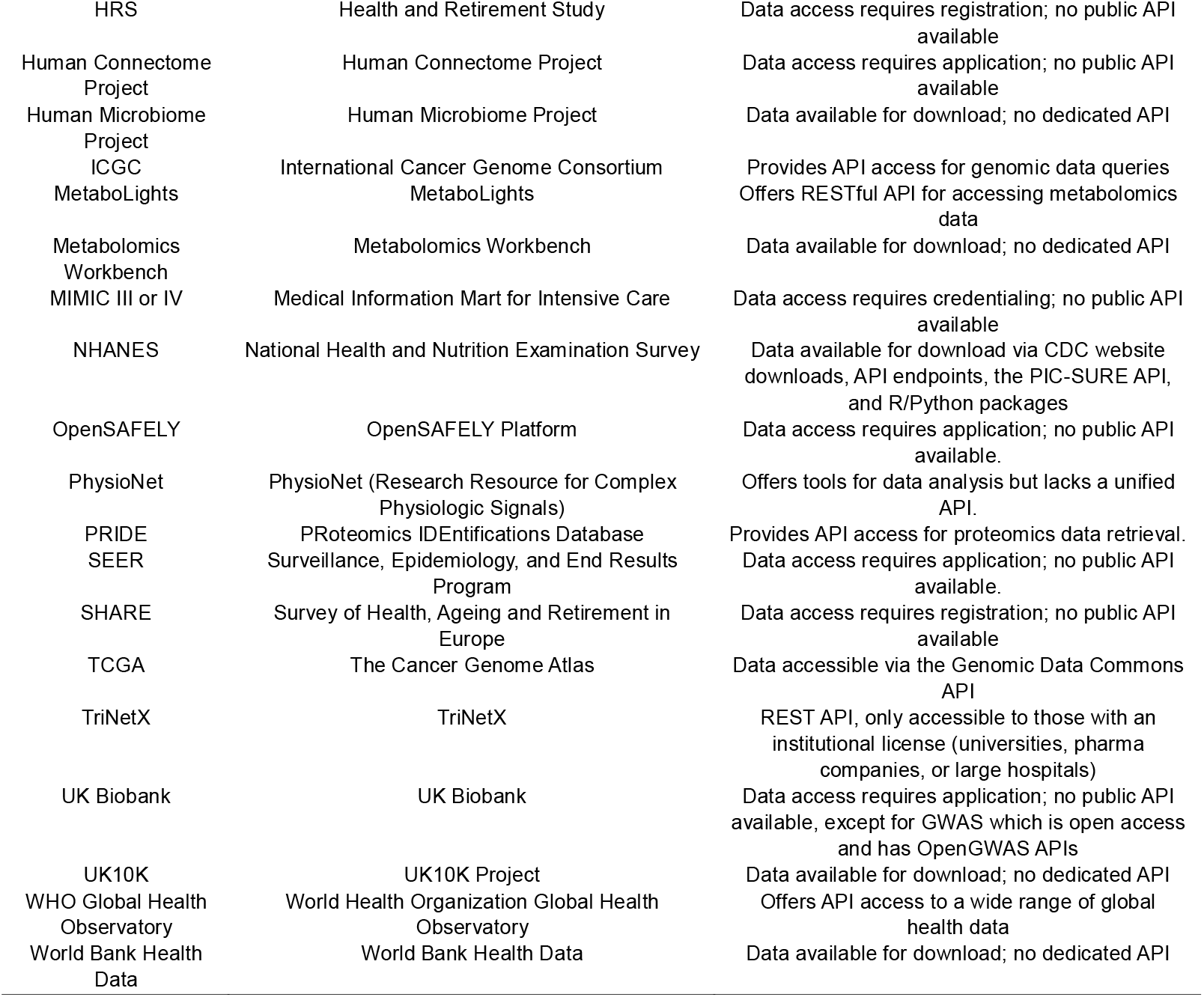
Data sources investigated, listed in alphabetic order.

The period from 2014 to 2022 was taken as a baseline largely undisturbed by the use of large language models (LLMs) and other forms of AI-assisted workflows, consistent with our previous work identifying an acceleration from late-2022 onwards. [4] 2023 to 2025 was considered to be the period substantially affected by technology-driven productivity gains from paper mills. To quantify the deviation from trend, for example where a data source might be experiencing a natural growth trend for non-exploitative reasons, forecasts were constructed using ARIMA (AutoRegressive Integrated Moving Averages). ARIMA is a time series forecasting algorithm with three components: autoregression (using past values), differencing (to account for trends and seasonality), and moving averages (to remove noise). The excess production of manuscripts was then taken as the difference between the observed production of manuscripts and the ARIMA forecasts. The ARIMA model used parameters of autoregressive order *p* = 1, degree of differencing *d* = 1 and moving average order *q* = 1. Confidence intervals were also constructed for the forecasts. ARIMA models were implemented in Python using the statsmodels library (version 0.14.4). [24]

Datasets where the publication rate in 2025 exceeded the 95% confidence interval for the ARIMA forecast were identified as potentially being exploited by new technologies – and therefore worthy of further investigation. “Genomics” was used as a control term in the OpenAlex database, as a more mature field where extensive prior work has been done by the research industry to reduce false discoveries and implausible conclusions. Secondary analyses were conducted to identify whether manuscript titles were becoming more homogenous / formulaic, and to identify datasets where there had been a geographic shift in the affiliations of last-named authors. For the identification of potentially formulaic titles, a simple count of increased frequency of words was conducted to test whether titles were becoming more homogenous. This was performed in Python using a count vectorizer function from the scikit-learn library (version 1.6.1). [25] For the geographic analysis, simple count of geographic origin was used together with fold changes of countries of origin between 2022 and 2025.

## Results

The fold-changes in publications for each data source for the period between 2022 and 2025 are summarised in Table 2. Of the searched datasets, four were excluded for having insufficient publication counts to support later modelling steps (UK10K, World Bank Health Data, OpenSAFELY, HiRID) and the All of Us Research Programme was excluded as its Researcher Workbench was only made available for data access in late 2020 to US researchers and for international researchers in late 2023. [26] Nine data sources (FinnGen, FAERS, NHANES, UK Biobank, Global Burden of Disease, CDC WONDER, TriNetX, CHARLS, and MIMIC III / IV) met the criterion of exceeding the 95% confidence interval for ARIMA forecasts. The median fold change was 1.2x, while the “genomics [title/abstract]” control search term produced a 2022-2025 fold change of 1.4x.

**Table 2.** Fold changes in publication numbers between 2022 and 2025 according to health data source, split between those with a change > median versus those with a change < median and ranked largest to smallest.

| Data sources with Fold Change > median (median = 1.2) | Fold Change | Data sources with Fold Change <= median (median = 1.2) | Fold Change |
| --- | --- | --- | --- |
| CDC WONDER | 17.4 | 1000 Genomes Project | 1.2 |
| TriNetX | 8.0 | BioBank Japan | 1.1 |
| All of Us Research Program | 6.9 | OpenSAFELY | 1.1 |
| FAERS | 5.6 | DHS | 1.1 |
| FinnGen | 5.5 | gnomAD | 1.1 |
| Global Burden of Disease | 3.2 | SEER | 1.1 |
| CHARLS | 2.9 | GTE<sub>x</sub> | 1.0 |
| HiRID | 2.5 | SHARE | 1.0 |
| NHANES | 2.4 | Human Connectome Project | 1.0 |
| UK Biobank | 2.3 | HRS | 1.0 |
| MIMIC-III or MIMIC-IV | 2.3 | PRIDE | 0.9 |
| ABCD Study | 2.1 | TCGA | 0.8 |
| WHO Global Health Observatory | 2.0 | PhysioNet | 0.7 |
| dbGaP | 1.9 | CCLE | 0.6 |
| MetaboLights | 1.9 | Human Microbiome Project | 0.6 |
| eICU Collaborative Research Database | 1.7 | ICGC | 0.5 |
| Metabolomics Workbench | 1.4 | UK10K | 0.4 |
| ADNI | 1.3 | World Bank Health Data | 0.0 |

The deviations from the ARIMA-estimated trend are shown in Figure 1.

**Figure 1:**
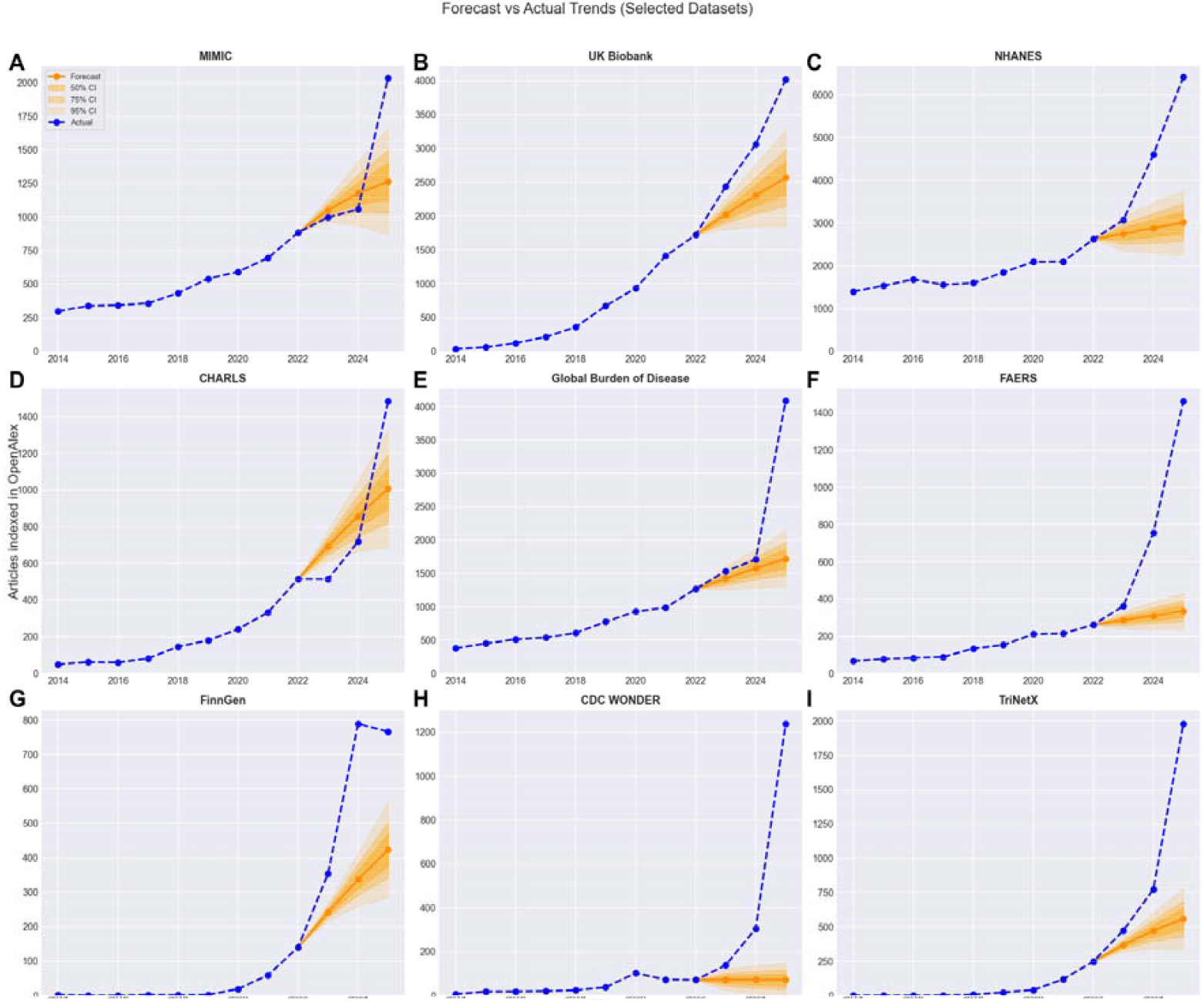
Actual publication counts compared with ARIMA forecasts, using 2014 to 2022 as the training period and 2023 to 2025 as the forecast period [A] MIMIC III / IV [B] UK Biobank [C] NHANES [D] CHARLS [E] Global Burden of Disease Study [F] FAERS [G] FinnGen [H] CDC WONDER, and [I] TriNetX. Numerical data underlying these figures are included in Supplementary Materials, Table S2

The data sources showing trend changes above the 95% confidence intervals of forecasts were then analysed for 2022 and 2025 to identify whether there was any change in titles of manuscripts (as a simple test for formulaic manuscript production). Nine datasets showed a sharp increase in certain tokens (words or phrases), as shown in Table 3, and this was especially marked for CDC WONDER (where 69% of titles commented on ‘trends’ in 2025, compared with 20% of titles in 2022. Analysis of the control search term ‘genomics’ showed that no tokens saw a change of more than 2.5% in frequency between 2022 and 2025, in contrast to the larger changes seen in the datasets suspected of exploitation. A list of the top 20 token changes is included in Supplementary Material, Table S3, including words which declined in usage.

**Table 3:** Increased incidence of tokens, measured by change in % of titles where each token was used.

| Dataset | Title tokens | 2022 % of titles featuring: | 2025 % of titles featuring: | Change % |
| --- | --- | --- | --- | --- |
| CDC WONDER | trends | 19.7 | 68.8 | 49.1 |
|  | cdc wonder | 7.0 | 52.7 | 45.6 |
|  | analysis | 18.3 | 54.3 | 36.0 |
|  | mortality | 52.1 | 88.0 | 35.9 |
|  | database | 5.6 | 27.3 | 21.7 |
| CHARLS | charls | 9.1 | 29.3 | 20.1 |
|  | association | 15.7 | 27.8 | 12.1 |
|  | index | 2.3 | 14.3 | 12.0 |
|  | risk | 11.1 | 22.5 | 11.5 |
|  | disease | 4.5 | 13.1 | 8.6 |
| FAERS | faers | 26.0 | 54.8 | 28.9 |
|  | analysis | 31.7 | 50.4 | 18.7 |
|  | database | 20.2 | 37.5 | 17.2 |
|  | world | 13.7 | 27.5 | 13.7 |
|  | safety | 7.3 | 20.2 | 12.9 |
| FinnGen | causal | 17.1 | 30.7 | 13.5 |
|  | insights | 0.0 | 7.8 | 7.8 |
|  | analysis | 13.6 | 20.6 | 7.1 |
|  | potential | 0.7 | 7.7 | 7.0 |
|  | relationship | 2.9 | 9.7 | 6.8 |
| Global Burden of Disease | analysis | 20.6 | 41.8 | 21.2 |
|  | global burden of disease | 25.1 | 44.7 | 19.6 |
|  | study | 31.9 | 46.5 | 14.6 |
|  | regional | 8.1 | 22.4 | 14.3 |
|  | trends | 13.3 | 27.4 | 14.1 |
| MIMIC III / IV | mimic-iv | 7.9 | 29.1 | 21.2 |
|  | database | 9.7 | 24.2 | 14.4 |
|  | retrospective | 9.4 | 22.8 | 13.4 |
|  | mortality | 18.3 | 31.1 | 12.7 |
|  | study | 12.4 | 24.3 | 11.8 |
| NHANES | nhanes | 21.5 | 43.4 | 21.9 |
|  | association | 26.2 | 41.4 | 15.2 |
|  | study | 14.0 | 26.6 | 12.6 |
|  | index | 5.5 | 16.9 | 11.4 |
|  | cross sectional | 8.2 | 18.5 | 10.3 |
| TriNetX | trinetx | 4.0 | 22.1 | 18.1 |
|  | study | 18.1 | 32.4 | 14.3 |
|  | cohort | 10.5 | 23.6 | 13.2 |
|  | matched | 4.4 | 16.5 | 12.1 |
|  | propensity | 4.8 | 15.7 | 10.8 |
| UK Biobank | risk | 28.5 | 32.5 | 4.0 |
|  | plasma | 0.3 | 3.8 | 3.5 |
|  | insights | 1.2 | 4.4 | 3.2 |
|  | cohort | 14.9 | 17.9 | 3.0 |
|  | prospective | 9.1 | 12.1 | 3.0 |

The nine data sources were then examined for changes to geographic origin, focusing on country of affiliation for the last-named author on each publication. The growth in publications is shown as a chloropleth in Figure 2. The largest change was for publications with affiliations located in China, which increased from 27% of publications indexed in the OpenAlex database in 2022 to 45% in 2025, or - in absolute terms - growth of 8,679 publications. Over the same period, the share of publications originating in the United States declined from 25% to 19%, as its annual publication count increased by 2,408 over the three years, with nearly half of this increase (n = 1,104) in publications analysing TriNetX.

**Figure 2:**
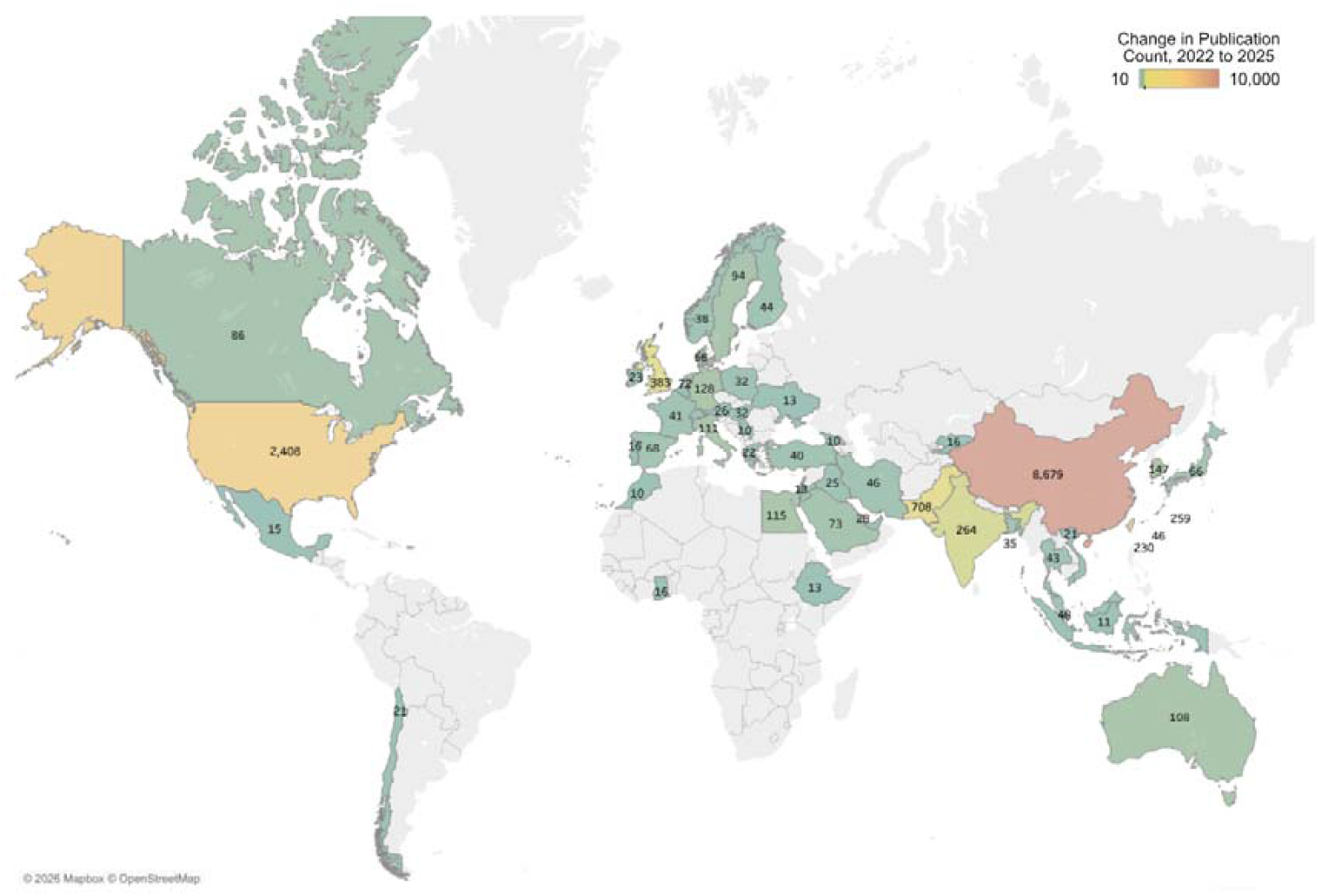
Chloropleth (geographical heatmap) of 2022 to 2025 change in annual publication count by country of author affiliations (fully counted), for the following nine datasets: CDC WONDER, CHARLS, FAERS, NHANES, UK Biobank, FinnGen, Global Burden of Disease Study, MIMIC III / IV, TriNetX. Filtered to only show countries with an increase in publications > 10 indexed in OpenAlex. Colour range is not linear and is centred at 200 publications. Data underlying this chloropleth are shown in Supplementary Materials, Table S4. As author affiliations are fully counted rather than fractionally, the total of affiliations does not sum to the number of publications.

Whilst the geographical split of growth in publications was very marked, it was not distributed evenly over the nine-fastest growing datasets. Seven saw a sharp increase in publications from China, based on full-attribution of affiliation (Table 4). This pattern was particularly evident for FAERS, where 66% of publications had an affiliation from China in 2025. Pakistan also showed a large change relative to its previous number, but this was focused largely within the CDC WONDER dataset. The rapid growth in publications using TriNetX was concentrated in the US.

**Table 4:** Table of fold changes for last-named author affiliations for the country seeing the largest absolute growth.

| Dataset | Country seeing largest Fold Change | 2022 affiliations from largest Fold Change country | 2025 affiliations from largest Fold Change country | Fold Change |
| --- | --- | --- | --- | --- |
| CDC WONDER | Pakistan | 1 | 553 | 553.0 |
| CHARLS | China | 442 | 1208 | 2.7 |
| FAERS | China | 102 | 843 | 8.3 |
| FinnGen | China | 76 | 569 | 7.5 |
| Global Burden of Disease Study | China | 427 | 2389 | 5.6 |
| MIMIC III / IV | China | 381 | 1017 | 2.7 |
| NHANES | China | 856 | 4043 | 4.7 |
| TriNetX | United States | 205 | 1309 | 6.4 |
| UK Biobank | China | 463 | 1596 | 3.4 |

## Discussion

Many approaches have been proposed for the identification of paper mill outputs or other problematic research practices, either reviewing individual papers [17] or seeking to identify concerning issues in overall publication trends (such as evidence of manipulated citation networks or retractions). [27,28] Here we use a scientometric approach to demonstrate that there has been rapid growth in numbers of publications since 2022 analysing open-source datasets, with nine datasets seeing both a break with previous trends that exceeded a 95% forecast confidence interval and signs of formulaic or templated research. This break in trend may also have been accelerated by the development of GenAI in late 2022, [29] but it is notable that the acceleration in publication rates differs between data sources. NHANES, FinnGen and the Global Burden of Disease Study experienced a break with trend in 2023, but FAERS publications saw an acceleration versus the previous trend only in 2024, with TriNetX and CDC WONDER seeing a larger acceleration in 2025. This may be suggestive of datasets being discovered at different times, albeit there are no obvious signs that datasets are being abandoned as other datasets start to be exploited.

Whilst comprehensive analysis of individual publications was beyond the scope of this work, many showed signs of titles becoming more formulaic, with ‘fingerprint’ methodologies, words and phrases consistent with previously described trends in the production of low quality manuscripts. Across the datasets, there was a strong trend towards reporting the name of the dataset used, and either ‘association’, ‘trends’ or ‘mortality’. This is consistent with our previous work which showed publications taking a well-described condition such as Type 2 Diabetes Mellitus, which is multifactorial in nature, and then analysing a single indicator or ratio for simple analysis and reporting the trend or association. Such simplistic studies fail to capture the complex and multifactorial nature of exposome-phenome associations. [30] The increased frequency of specific tokens and methodologies (for example the majority of the studies investigating CDC WONDER used Joinpoint software and the same workflow) is suggestive of a template-driven improvement in productivity and output, possibly also using Generative AI. [31]

Coincident with these changes there was also a dramatic shift in geographic origin of corresponding (last-named) author affiliations for the 9 most affected databases. The absolute increase for China between 2022 and 2025 (8679 publications) dwarfed that for the United States (2408), with the next three largest increases from Pakistan (708), the UK (383) and India (264). Authors with affiliations to Chinese hospitals have been associated with high retraction rates of papers in various fields, [32] and researchers from China are also subject to different incentives, [33] as are scientists in lower-middle income countries, [34] often due to difficulties in undertaking and publishing research and a lack of institutional support. Furthermore, the Chinese supreme court has recently issued guidance on paper mills and on scientific fraud, calling for lower courts to crack down on the mass-produced academic paper industry. [35]

Overall, of the datasets showing an acceleration in publication rates, the additional evidence of formulaic titles and a geographic shift in authorship is strongly suggestive of a change in publication patterns for nine of the datasets analysed in this work. These are FAERS, NHANES, FinnGen, UK Biobank, the Global Burden of Disease Study, CHARLS, MIMIC III / IV, TriNetX, and CDC WONDER. The rapid changes over the four-year period may well be due to paper mill exploitation. Eight of the nine datasets also share open access elements, often with APIs that facilitate highly productive workflows. For the UK Biobank and FinnGen, open-access GWAS data were typically used, [36] NHANES has a well developed API system, [37] FAERS research is facilitated through OpenVigil, [38] and whilst the Global Burden of Disease Study does not have an API the data are freely available for download. We have written previously on 2SMR and association templates, but drug safety is a newly exploited type of data. FAERS itself is a valuable asset in a field that has not always had full data transparency, [39] but is a voluntary reporting system and so cannot provide estimates of incidence, cannot analyse causality, and additionally has considerable potential for bias based on physician or public preferences. [40] Misleading publications based on such drug safety reports pose a particular risk to the public, either by amplifying unnecessary concerns or underplaying risks. TriNetX presents a different profile, with many of the publications targeted at conferences and a US-centric production model focused on medical schools. This illustrates that the demand for mass-produced manuscripts is not an issue related solely to the Global South.

Such problematic outputs present clear harms. Taken individually, the papers we find are simply not very interesting (they are formulaic and repeat the same analyses across thousands of variables), but taken together they become misleading through the introduction of false discoveries, dilution of genuine findings in the scientific literature, and reduced credibility for high quality data sources, An additional issue is that many GenAI models are trained on - and learning from - scientific literature. With the inputs for learning potentially becoming corrupted, these models will propagate false science. For the nine most problematic datasets identified here, the excess publications above trend (around twelve thousand) represented 50% of the 2025 output, a significant proportion to be potentially misleading.

We and others have previously written on measures to mitigate against formulaic manuscript production, focusing on more frequent use of desk rejections by journals to reduce burdens on peer reviewers, dedicated statistical reviewers, [41] use of application numbers by data providers and more effective post-publication correction. [42,43] Others have also written on the need for effective tools to identify problematic manuscripts such as the recently-proposed GRABDROP checklist, [44] and we suggest that increased awareness of datasets currently experiencing worrying trends and key phrases associated with formulaic outputs will assist in this process. [7,27,45] It is, however, important to stress that the threat to scientific integrity identified here presents risks to the wider goals of Open Science and compliance with the FAIR Guiding Principles. [19,46] The factors that drive compliance with these principles also make such datasets vulnerable to exploitation. To preserve the overall goal of Open Science and FAIR compliance, we believe that unrestricted open access to AI-ready data may not be the best option for all data resources. For some, approved access and / or pre-registration may offer safeguards against exploitative data dredging. A more radical solution given the ease of analysing simple relationships such as ‘The association of [Predictor A] with [Outcome B] in [Population C] using a cross-sectional national dataset [Open Access Dataset D]’ would be for the scientific community to deem such results as effectively already available online, and so not suitable for publication.

A number of limitations should be stressed in this work, the most significant of which is that this is a scientometric analysis, rather than a comprehensive review of individual papers. For example, GenAI has the potential to generate formulaic low-quality outputs, sometimes referred to in other fields as “AI slop”. [47] It is challenging from the outside to tell the difference between deliberate, coordinated mass production by paper mills and uncoordinated mass production by individuals facilitated by large language models, and this is a limitation of this research. A second limitation is that accelerations in publication may be due to good reasons. For example, the UK Biobank has made a number of new datasets available over time. Third, this analysis is retrospective, and future behaviour may change in response to recent publisher policy changes to restrict submissions dealing with open access datasets, such as NHANES. [48,49] Given the adaptive and adversarial nature of unethical actors, we expect both scientometric analyses and targeted investigations to form part of an ongoing effort to protect research integrity and the principles of Open Science, especially given that paper mills will adapt their strategies as existing approaches are brought to light. [50,51] It should also be noted that in this work we have not integrated resources such as the Problematic Paper Screener; this may be an opportunity for future work, albeit these traditional flags (for example for the detection of tortured phrases) [52] are not typically found in this type of formulaic manuscript, possibly due to adaptive changes by paper mills. Finally, it should be noted that scientometric analysis does not substitute for existing methods of detecting problematic manuscripts, such as citation networks, evidence of image manipulation or text similarities, as well as the backstop of peer review and editorial assessment. [17,53–55] Nonetheless, the fact that this work reviews accepted manuscripts provides evidence that existing tools used by publishers may be inadequate for new challenges posed by paper mill, [18,56,57] especially when post-publication corrections / retractions are often slow, or may not happen at all. [58] Furthermore, even retracted articles can contaminate evidence syntheses, and the sheer weight of publications flagged in this work is suggestive that these negative downstream impacts are likely to increase. [59,60]

In conclusion, our scientometric analysis highlights significant concerns about research integrity, particularly in light of previously documented associations between institutional pressures and increased retraction rates and the risk of paper mills employing new AI-supported workflows to produce manuscripts on an industrial scale. This growth poses a direct threat to the core principles of scientific rigour, and those of Open Science and FAIR data practices, given that the accessibility and interoperability designed to facilitate legitimate research also enables exploitation. To mitigate these risks and safeguard scientific integrity, we advocate for controlled data-access models paired with mandatory pre-registration of analyses, rather than unrestricted open access. This balanced approach is essential to preserving the intended benefits of Open Science while preventing its misuse, ensuring that scientific advancement continues to be reliable, reproducible, and trustworthy.

## Supplementary Materials

Table S1: Search Strings. Table S2: Annual Data. Table S3: 20 tokens seeing the largest change in titles between 2022 and 2025, by ‘at risk’ dataset. Table S4: Matrix of change in publication count by country for the combined nine ‘at risk’ datasets

## Author Contributions

Matt Spick: Conceptualization, Methodology, Software, Formal analysis, Investigation, Writing—original draft preparation, Visualization, Project administration. Anthony Onoja: Data curation. Charlie Harrison: Formal analysis, Investigation, Writing—review and editing. Stefan Stender: Methodology, Writing—review and editing. Jennifer Byrne: Methodology, Writing—review and editing. Nophar Geifman: Conceptualization, Resources, Writing—review and editing, Supervision. All authors have read and agreed to the published version of the manuscript.

## Data Availability Statement

All data used in the preparation of this manuscript are included in Supplementary Materials. All code used in this work employed standard Python libraries without any modifications.

## Acknowledgments

The authors wish to acknowledge the wider support of the United2Act network as well as the AIBIO-UK network

